# Identification of Intimate Partner Violence from Free Text Descriptions in Social Media

**DOI:** 10.1101/2021.12.15.21267694

**Authors:** Phan Trinh Ha, Rhea D’Silva, Ethan Chen, Mehmet Koyutürk, Günnur Karakurt

## Abstract

Intimate Partner Violence (IPV) is a significant public health problem that adversely affects the well-being of victims. IPV is often under-reported and non-physical forms of violence may not be recognized as IPV, even by victims. With the increasing popularity of social media and due to the anonymity provided by some of these platforms, people feel comfortable sharing descriptions of their relationship problems in social media. The content generated in these platforms can be useful in identifying IPV and characterizing the prevalence, causes, consequences, and correlates of IPV in broad populations. However, these descriptions are in the form of free text and no corpus of labeled data is available to perform large-scale computational and statistical analyses.

Here, we use data from established questionnaires that are used to collect self-report data on IPV to train machine learning models to predict IPV from free text. Using Universal Sentence Encoder (USE) along with multiple machine learning algorithms (Random Forest, SVM, Logistic Regression, Naïve Bayes), we develop DetectIPV, a tool for detecting IPV in free text. Using DetectIPV, we comprehensively characterize the predictability of different types of violence (Physical Abuse, Emotional Abuse, Sexual Abuse) from free text. Our results show that a general model that is trained using examples of all violence types can identify IPV from free text with area under the ROC curve (AUROC) 89%. We also train type-specific models and observe that Physical Abuse can be identified with greatest accuracy (AUROC 98%), while Sexual Abuse can be identified with high precision but relatively low recall. While our results indicate that the prediction of Emotional Abuse is the most challenging, DetectIPV can identify Emotional Abuse with AUROC above 80%. These results establish DetectIPV as a tool that can be used to reliably detect IPV in the context of various applications, ranging from flagging social media posts to detecting IPV in large text corpuses for research purposes. DetectIPV is available as a web service at https://ipvlab.case.edu/ipvdetect/.

## 1 Introduction

Intimate Partner Violence (IPV) is a public health problem that affects millions of individuals worldwide. IPV is considered as any behavior by a current or former partner that causes harm to those involved in the relationship ^1^. IPV includes physical, sexual, and emotional forms of violence and aggressive acts ^2^. Physical violence is defined as the use of physical force and actions to inflict physical harm in a partner. These actions include, but are not limited to, slapping, pushing, kicking, punching, cutting, burning, and using weapons. Sexual violence can be defined as forcing or attempting to force a partner to take part in a sex act, sexual touching or a non-physical sexual event when the partner does not or cannot consent. Emotional violence is assessed by measuring harms to emotional well being.

According to the World Health Organization, it is estimated that on average 30% of women experience IPV globally ^3^. Based on the National Intimate Partner and Sexual Violence Survey, about 1 in 4 women as well as 1 in 10 men reported experiencing some form of IPV in the US ^2 4^. Unfortunately, IPV is also among the leading causes of death among younger women ^1 5^. Statistics indicate that about one third of all women murdered in the United States are killed by their partner ^6^.

IPV causes numerous physical, psychological, sexual, and emotional trauma. Past research showed that victims of IPV are more likely to suffer from physical, mental, and sexual health issues in acute or chronic manner, than non-victims ^7 8^. Health problems ranging from minor cuts to broken bones, injuries, disability and other severe health consequences due to IPV are frequently reported by the victims ^9^. Furthermore, mental health issues such as post-traumatic stress disorder, clinical depression, and suicide attempts are highly prevalent among the victims of IPV ^7 10 3^. Recent research also indicates that emotional abuse caused by degradation, intimidation, and control, is highly prevalent in intimate relationships and co-occur frequently with physical violence ^11^. For these reasons, effective identification of IPV in various settings can be useful in raising awareness, providing help to victims, and generating data for research to further understand the causes, consequences, and treatment of IPV.

### Detection and Measurement of IPV

Detecting IPV in various settings can be challenging. Over time, various measurement approaches have been used by the scientific community to assess the nature and extend of IPV in relationships. These approaches heavily relied on measuring frequencies and occurrence rates of specific violent behaviors. The most widely used scale to assess both the perpetration and victimization is the Conflict Tactics Scale (CTS) ^12 13^. CTS measures frequencies of perpetration and victimization of physical, emotional, and sexual violence that happened in the last year. Other scales such as the Abuse Behavior Inventory (ABI) use variations of the CTS as behavioral checklists that define abuse acts through specific behaviors.

Emotional abuse is a complex construct that includes various emotional tactics by the abuser for continuous emotional mistreatment of a person. It can be defined as the use of verbal and non-verbal communication with the intent to harm another person mentally or emotionally and/or to exert control over another person. Acts demonstrating emotional abuse are thought to be along a spectrum, with deliberate scaring, humiliation, and isolation as well as threats to an individual’s safety. Due to the complexity and the spectrum of emotional abuse, as well as its nonverbal components, researchers and clinicians face challenges in consistently assessing emotional abuse. Widely used emotional abuse scales such as the Psychological Maltreatment Inventory (PMI) ^14^ and the Women’s Experience with Battering (WEB) scale ^15^ define emotional abuse also through specific behaviors.

Measuring sexual violence in intimate relationships also faces many challenges as the definition is often viewed to be synonymous with rape. However sexual violence consists of a spectrum of behaviors such as sexual demands that make partners uncomfortable, pressuring, controlling, and manipulating to obtain sex and degradation of partners. To understand the extent of the problem in intimate relationships, scales such as Sexual Experiences Survey (SES) ask questions regarding sexual experiences and assign participants into categories of non-victimized, sexually coerced, abused, or assaulted based on the severity of sexual violence ^16^.

### Use of Social Media to Characterize Public Health Problems

With the growing popularity of social media platforms, the amount data generated from user posts and activities along with the availability of tools to collect user information is more than ever before. As a result, patterns and indicators of public health problems can be extracted from the social media, in an attempt to better characterize these problems, assess their prevalence, and identify their association with other issues. In recent literature, there have been many applications of using Social Media to characterize public health problems from large amounts of data generated from social media platforms ^17^. In relation to problems of substance abuse, social media platforms such as Instagram and Twitter provide a multitude of data formats to characterize, in the form of both text and images posted by users on social media platforms ^18^. Images can be used in conjunction with image tagging tools to generate description tags for user’s posts and can then be used for inferring a local region’s public health problem. ^19^. With the abundance of free floating text in users post, text data is also commonly used in conjunction with domain expertise for encoding to characterize behaviors ^17 18^.

### Challenges in Predicting IPV from Social Media

In identifying public health problems, sets of keywords can be used as identifiers. However, IPV can become more context-dependent and could be described by victims or perpetrators in different ways. In this case, use of keywords alone can overlook cases of IPV, and as such should require a use of a higher level representation to describe. Thus more sophisticated machine learning and natural language processing (NLP) algorithms can be potentially more effective in detecting IPV in social media data. However, there are no existing labelled datasets with regards to IPV to train sophisticated machine learning algorithms. In particular, there is no readily available corpus of accounts of victims of IPV cases. As a result, manual curation and labelling is required in order to generate datasets for training and testing machine learning models to predict IPV in social media data.

### Our Solution and Contributions

To overcome the challenges posed by data, identification, and model building, we present a comprehensive framework with the following components:

- We use established questionnaires from the literature as training data. This provides a corpus for training machine learning models in addition to the readily trained models that we build and validate in this study.
- We use natural language embedding based text embedding models to enable training of standard representation learning algorithms on free text. Importantly, the embeddings can be computed based on larger corpuses that are not necessarily labeled.
- We use Reddit’s `r/relationship_advice` platform as our data source from social media for testing and validation. For this purpose, we use a list of keywords curated by our domain experts to generate a set of candidate posts from `r/relationship_advice`, and use manual labeling to characterize the existence/description of violence in each sentence per post. We also randomly select posts/sentences from other subreddits to generate different types of negatively labeled samples. Besides serving as test data for the validation of our machine learning models, these labeled data also provide a valuable resource for data-driven discovery using other computational approaches.
- We use a variety of machine learning algorithms, collectively named DetectIPV, to comprehensively characterize the predictability of violence. Using this framework, we develop general as well as type-specific violence models to characterize different types of IPV.
- To make DetectIPV available to the public, clinicians, and the scientific community, we release DetectIPV as a web service with an easy-to-use interface and visualizations that allow exploratory analyses. The web service is available at http://ipvlab.case.edu/ipvdetect/.

## 2 Methods

### 2.1 Study Design

The purpose of our study is to identify intimate partner violence (IPV) from free text. In our computational experiments, we focus on social media posts, but the text can also be in the form of scientific articles, free-floating notes from clinicians, and case interviews. While the posts can be (and usually are) composed of multiple sentences, we consider sentences as units of analysis as this simplifies the task of learning.

#### Labeling of Sentences

IPV can include multiple types of violence including physical, emotional, and sexual abuse. To capture the specificity of types of violence, we categorize sentences using five different labels:

- **General Violence (GV):** The sentence describes a situation that involves IPV.
- **Physical Abuse (PA):** The sentence describes a situation that involves physical abuse. PA is defined as any form of intentional force with the potential for causing death, disability, injury or harm. It includes behaviors when a person hurts or tries to hurt a partner by hitting, kicking, or using another type of physical force. It can also be acts of physical assault, use of weapons and threats of assault by a partner.
- **Emotional Abuse (EA):** The sentence describes a situation that involves emotional abuse. EA involves the continual emotional mistreatment of a person. It is the use of verbal and non-verbal communication with the intent to harm another person mentally or emotionally and/or to exert control over another person. Emotional abuse can involve deliberately trying to scare, humiliate, isolate, or ignore a person. It refers to behaviors that harm a person’s self-worth or emotional well-being. Emotional abuse is the form of abuse that is not physical, rather emotional. This can include the diminishing of self-worth or identity, lack of independence, neglect, and extends to verbal abuse.
- **Sexual Abuse (SA):** The sentence describes a situation that involves sexual abuse. Sexual abuse/violence is defined as any unwanted sexual activity, making unwanted sexual advances or abusing lack (or incapability) of consent. It is any sexual act, attempt to obtain a sexual act or other act directly against a person’s sexuality using coercion, by any person regardless of their relationship to the victim in any setting. Sexual violence not only includes abuse/assault or rape but also includes sexual exploitation, sexual harassment, stalking, and cyber harassment. For the purpose of this research, we focus on experiences between two people in a relationship.
- **Negative (NE):** The sentence does not describe a situation that involves IPV. We consider two types of sentences that are considered as negative: (i) *Unrelated Negatives* are sentences that are irrelevant to relationships or violent situations. (ii) *Related Negatives* describe situations that involve relationship problems but do not include violence or abuse, or describe healthy and/or happy relational interactions.

Physical Abuse, Emotional Abuse, and Sexual Abuse are subcategories of General Violence that are not mutually exclusive. In other words, if a sentence is labeled GV, then it needs to be also labeled by ***at least one of*** PA, EA, or SA. If a sentence is labeled NE, then it cannot be labeled with any of PA, EA, or SA.

#### Machine Learning Task

We set up the machine learning task as follows: Given a sentence, decide whether it describes a citation that involves IPV, in the form of one or more of physical, emotional, or sexual violence. In our framework, we train “General Violence” models to distinguish the category GV from the category NE, as well as “Type-Specific” models to distinguish each of PA, EA, and SA from NE. We use the distinction between unrelated and related negatives to characterize what the models are learning (e.g., are they learning how to distinguish relationship problems from irrelevant situations or are they learning to distinguish violent situations in relationships from non-violent situations in relationships?)

#### Computational Workflow

The workflow of our framework is shown in Figure 1. For training, we obtain items from selfreport questionnaires that are used in the literature to identify, quantify, or characterize violence in relationships. We then use Universal Sentence Encoder (USE) to generate vector-space embeddings of these questionnaire items. Subsequently, we use the embeddings and labels of the questionnaire items to train predictive models In parallel, we perform literature review to identify keywords that are associated with IPV (“Terms of Interest”). We use these keywords to identify social media posts that are potentially associated with IPV (“Top Posts”). We manually investigate these posts to label each sentence in the post as positive or negative, also annotating the positive sentences with the type of violence. These sentences form our test data. We generate vector-space embeddings for test sentences using Universal Sentence Encoder (USE) as well. Using these embeddings and the machine learning models trained on questionnaire items, we predict the labels of the test sentences and evaluate the predictive accuracy of the models.

**Figure 1:**
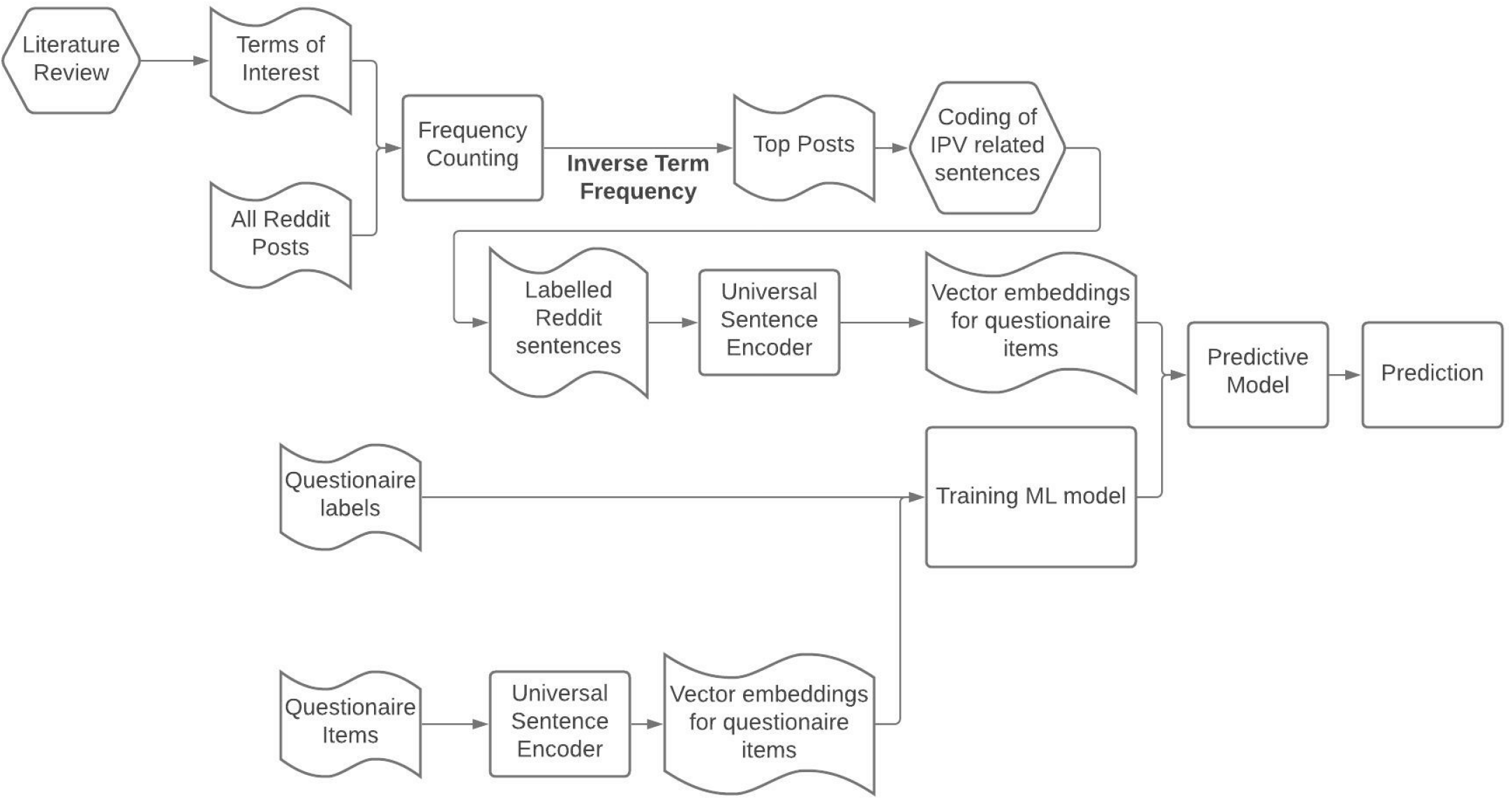
The workflow of the machine learning framework for predicting IPV from social media posts. Boxes with curved sides show data (input, intermediate, or output), honeycombs show human input (expert review, literature search coding), and rectangles show algorithms/tools/processing.

### 2.2 Dataset Retrieval and Curation

#### Training Data: Questionnaires

For training, we utilize self-report questionnaires that are used in the literature to assess conflict and/or violence in a relationship. The items in these questionnaires represent descriptions of a wide range of violent interactions that are commonly accepted as examples of IPV by the scientific community. Use of questionnaires for training serves two purposes: (i) the training data is reliable (based on scientific literature), (ii) all the data obtained from social media can be for testing. This is important as social media data is not readily labeled, and thus requires a large amount of manual work for labeling. The questionnaires that are used to obtain training data are shown in Table 1.

**Table 1:** Questionnaires and scales used as training examples. Top: Positive examples, Bottom: Negative examples.

| Physical Violence | Emotional Violence | Sexual Violence |
| --- | --- | --- |
| Abusive Behavior Inventory<sup>20</sup> | Abusive Behavior Inventory<sup>28</sup> | Measure of Wife Abuse<sup>23</sup> |
| Composite Abuse Scale<sup>21 22</sup> | Composite Abuse Scale<sup>21 22</sup> | National Women's Study (NWS) and National Violence Against Women Survey (NVAWS)<sup>37 38 39</sup> |
| Measure of Wife Abuse<sup>23</sup> | Index of Psychological Abuse<sup>29 30 31</sup> | Revised Conflict Tactic Scale<sup>13</sup> |
| Partner Abuse Scale - Physical<sup>24</sup> | Measure of Wife Abuse<sup>23</sup> | Severity of Violence Against Women Scale (SVAWS)<sup>27</sup> |
| Revised Conflict Tactic Scale<sup>13</sup> | Multidimensional Measure of Emotional Abuse<sup>32</sup> | Sexual Experiences Survey (SES) - Victimization Version<sup>40 16 41</sup> |
| Safe Dates - Physical Violence Victimization<sup>25 26</sup> | Partner Abuse Scale - Non-physical<sup>33</sup> | Sexual Victimization of College Women<sup>42 43</sup> |
| Severity of Violence Against Women Scale (SVAWS)<sup>27</sup> | Psychological Maltreatment of Women Inventory (PMWI)<sup>14 34</sup> |  |
|  | Psychological Maltreatment of Women Inventory (PMWI)—Short Form<sup>34</sup> |  |
|  | Revised Conflict Tactics Scales (CTS-2)<sup>13</sup> |  |
|  | Safe Dates — Psychological Abuse Victimization<sup>25 26</sup> |  |
|  | Women's Experiences with Battering (WEB)<sup>15 35 36</sup> |  |
| Related Negatives | Unrelated Negatives |  |
| Revised Dyadic Adjustment Scale<sup>44</sup> | Marlowe-Crowne Social Desirability Scale 13-Item Short Form<sup>55</sup> |  |
| Coping Inventory for Stressful Situations<sup>45</sup> | The Big Five Inventory Personality Test<sup>56</sup> |  |
| Experiences in Close Relationships Scale<sup>46</sup> | The Patient Satisfaction Questionnaire Short Form<sup>57</sup> |  |
| Dominance Scale<sup>47</sup> | Multifactor Leadership Questionnaire<sup>58</sup> |  |
| Short Form Health Survey (SF-36)<sup>48</sup> | Academic Progress Questionnaire<sup>59</sup> |  |
| Dyadic Coping Inventory<sup>49</sup> | Student Engagement in Schools Questionnaire<sup>60</sup> |  |
| Marital Burnout<sup>50</sup> | College Students Financial Literacy Survey (CSFLS)<sup>61</sup> |  |
| Trauma Symptom Checklist<sup>51</sup> | Driver's adaptive glance behavior<sup>62</sup> |  |
| The Positive and Negative Affect Schedule<sup>52</sup> | Creature of Habit Scale<sup>63</sup> |  |
| Interpersonal Reactivity Index<sup>53</sup> | Lifestyle Questionnaire (British Heart Foundation)<sup>64</sup> |  |
| Brief Symptom Inventory<sup>54</sup> | Assessment of Leisure Time Physical Activities<sup>65</sup> |  |
|  | Student Teacher Relationship Survey<sup>66</sup> |  |
|  | Multifactorial Memory Questionnaire<sup>67</sup> |  |
|  | Conceptualizing Leadership Questionnaire<sup>68</sup> |  |

Positively labeled samples for training data (GV, PA, EA, SA) are items from the CDC’s compendium of assessment tools for IPV ^69^. The items representing related negatives represent sentences containing words that also exist in IPV-associated sentences. For example, the sentence `“My partner told me that I am worthless”` describes Emotional Abuse, but the word `“worthless”` can be contained in the sentence `“I feel worthless”`, which is not an indicator for IPV. The unrelated negatives are questionnaire items that are irrelevant to IPV and other relationship issues, taken from multiple unrelated topic questionnaires.

Overall, the training data includes 355 positively labeled and 707 negatively labeled sentences. Among the positively labeled sentences, 126 are labeled as Physical Abuse, 198 are labeled as Emotional Abuse, and 31 are labeled as Sexual Abuse. Among the negatively labeled sentences, 410 are unrelated negatives while 297 are related negatives. All sentences that are used as positive and negative training samples are provided in Supp. Table 1.

#### Test Data: Social Media Posts

The test data is collected from the subreddit `r/relationship_advice` from the social media site Reddit, from posts that were created between January 2019 and July 2020, using PushShift’s API. After processing, This dataset contains 116 posts composed of a total of 326 sentences.

To identify posts that are potentially related to IPV, we use a word-frequency based approach. For this purpose, we manually curate a list of 119 terms that are potentially related to IPV. These keywords are listed in Supp. Table 2. Using these terms, we generated a set of candidate posts that can potentially describe IPV situations. We then manually evaluate these posts to identify sentences that describe situations involving IPV.

The process of generating the set of candidate posts is as follows:

- Lemmatize all words in posts and terms so that all words are simplified to their basic form (e.g., both `punching` and punches are reduced to `punch`).
- Compute the frequency of all 119 terms from all collected posts. Let *f* (*t*) denote the frequency of term *t* in the corpus.
- Compute the score for each post D as *score* 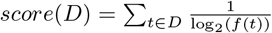 Use of inverse term frequency ensures diversification of the posts, i.e. posts with uncommon keywords are not overwhelmed by posts with more common keywords.
- Rank the posts in decreasing order of *score*(*D*). For manual evaluation, we focus on the 90 posts with highest scores. We call these posts candidate posts.

Each candidate post is then evaluated and labeled by two experts, sentence by sentence, to match our machine learning framework. For this purpose, we develop a detailed manual for the operational definitions of different types of IPV, and their underlying concepts. These operationalizations with multiple examples helps the team identify the presence of different types of violence in qualitative descriptions of relational experiences. Team members are trained in recognizing the signs of the relational problems over an hour long weekly meetings for 6 months.

During the process of evaluation and labeling, our team found that sentences with IPV can contain more than one type of IPV. For example, the sentence `“No matter where we were, when she was angry she would not hesitate to verbally and physically abuse me”`, contains both Physical Abuse and Emotional Abuse. Consequently, in our labeling, we allow a positively-labeled sentence to be assigned more than one type of IPV (PA, EA, SA). As a result, we obtain 326 positively-labeled test sentences, of which 74 are labeled as PA, 257 are labeled as EA, and 25 are labeled as SA (with some sentences containing overlaps).

We obtain “Unrelated negative” test sentences by extracting posts from an unrelated subreddit. Namely, we collect 196 posts on `r/changemyview` and select a random sentence from each post. For “Related negative” test sentences, we randomly selecte sentences from the original corpus we obtained from `r/relationship_advice`, which are verified by the curation team to ensure that these sentences are relationship related, but do not describe any IPV-related situations. As a result, we obtain 292 negatively labeled test sentences, of which 196 are considered unrelated negatives and 96 are considered related negatives. The list of positive and negative test sentences is provided in Supplemental Table 3.

The resulting number of training and test sentences and the distribution of their labels are shown in Table 2.

**Table 2:** Training and testing data used in our computational experiments. Each entry shows the number of positive and negative examples of the respective type in the training and testing data. The total number of positive examples is less than the sum of the number of examples in each violence type, since a single sentence can be annotated with multiple violence types.

| Dataset | Positive Examples |  |  |  | Negative Examples |  |  |
| --- | --- | --- | --- | --- | --- | --- | --- |
|  | Physical | Emotional | Sexual | Total | Related | Unrelated | Total |
| Training: Questionnaire Items | 126 | 198 | 31 | 355 | 297 | 410 | 707 |
| Testing: Sentences from Reddit Posts | 74 | 257 | 25 | 326 | 96 | 196 | 292 |

### 2.3 Model Building

#### Computation of Sentence Embeddings

As it is difficult to interpret unstructured data, transformation of unstructured data into a more structured format is usually desirable. This is often accomplished by embedding “unstructured” samples into a low-dimensional vector space ^70;71^. In the context of natural language processing, there are algorithms for embedding words, sentences, or documents. Since our focus here is on sentences, we use Universal Sentence Encoder (USE) ^72^, a sentence encoder with documented success in various applications ^73 74 75^. Given a sentence, USE uses neural network architectures that are pre-trained on large corpuses of text to identify latent dimensions that are descriptive of the variance in these corpuses. This is particularly useful in our application as it removes the requirement for abundant data to compute robust and descriptive embeddings.

USE works on a per-sentence basis, and is designed specifically for transfer learning, where the model’s output can be reused as part of another task. USE takes in a string sentence and output a fixed 512-dimensional vector representing the sentence. These embeddings take into account sentence semantics, such that sentences with similar meanings are closer to each other in the embedding space. Proximity in the embedding space is quantified in terms of the angular distance between the embeddings *u* and *v* of a pair of sentences:

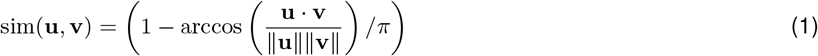

USE has two different model architectures, one with Deep Averaging Network (DAN) ^76^ and the other with a Transformer ^77^. In our application, we use a pre-trained DAN based encoder. In this architecture, the output token embeddings, which are dense vectors that correspond to a token, are averaged. USE implements this architecture by generating embeddings from each word and bi-gram token, which are then passed through a deep feed-forward network to compute the sentence embedding as a non-linear combination of these word and bi-gram embeddings.

#### Feature Selection

The embeddings provided by USE do not take into account our training data and map the sentences to a fixed 512-dimensional space. For this reason, it is potentially useful to order these dimensions in terms of their discriminative potential based on the training data and perform feature selection. We take a filtering-based approach to feature selection and rank the sentences according to their confusion in distinguishing positive training examples from negative training examples. For this purpose, we formulate a penalty function that assesses each dimension’s ability to separate positive examples from negative examples in the training data: Letting 𝒟 denote the set of all dimensions returned by USE and 𝒩 denote a given set of negative examples we define the *separability* of a pair *p*_*i*_ and *p*_*j*_ of positive samples with respect to a given set of dimensions *D* ⊆ 𝒟 and a as:

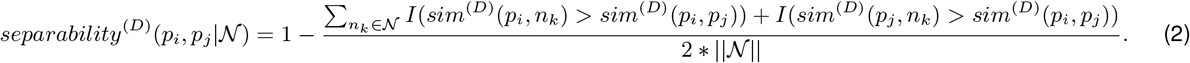

Here *sim*^(*D*)^(*u, v*) denotes the similarity between the embeddings of two samples in the space represented by *D*. and *I* denotes indicator function.

Then, for a given set of positive examples 𝒫 and negative examples 𝒩, we quantify the separability of 𝒫 and with 𝒩 respect to *D* as:

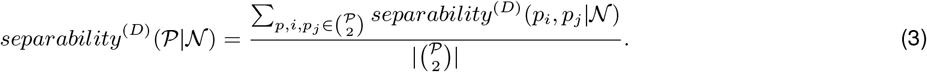

With these measures in hand, we compute the separability of all positive sentences and negative sentences in the training data with respect to each USE dimension *d* ∈ 𝒟. We order the USE dimensions in decreasing order of *separability*^(*{d}*)^(𝒫| 𝒩), and compute the *separability*^(*D*)^(𝒫|𝒩) where *D* is a growing set of dimensions with each dimension added in order of individual separability. We then select the set of dimensions where separability on training data peaks and use those dimensions to train the models. Comprehensive results on feature selection and the effect of the number of embedding dimensions on predictive performance are shown in Section 3.4.

#### Training Machine Learning Models

To build a predictive model using USE embeddings of the sentences from questionnaires, we use four different machine learning algorithms. These algorithms are Logistic Regression, Naive Bayes, SVM and Random Forest. We then evaluate the ability of the model in detecting IPV in sentences extracted from Reddit posts. We pick these algorithms as they have been successfully applied in the past with similar applications, including mental statement assessment in social media ^78^. We use social media data from Reddit for testing to investigate how well the models learned from the questionnaires generalize to practically relevant text data.

#### Type-Specific vs. General Violence Models

In addition to training a general model to identify intimate partner violence in general, we also train type-specific models to identify specific types of IPV, namely Physical Abuse (PA), Emotional Abuse (EA), and Sexual Abuse (SA). While the training for the general violence model incorporates all positively labeled samples, we train the type-specific models by considering the positively labeled samples that are labeled with the respective subtype. As negative training samples, both the general and type-specific models use the negatively-labeled samples. For each sentence, each trained model produces a confidence value indicating the likelihood of the sample belonging to a type of IPV. We use these confidence values to visualize the sentences in the space of the three types of IPV and explore the relationship between each subcategory of IPV.

## 3 Results

In all of our computational experiments, we use questionnaire data for training and social media data for testing. Our computational model uses sentences as units of analysis. The number of sentences in the training and testing datasets, and their distribution into violence subtypes and types of negatives are shown in Table 2.

### 3.1 Predictive Performance of the General Violence Model

We first assess the predictive performance of the General Violence Model using four different classification algorithms. In this model, sentences with violence in the training (questionnaire) data are labeled as “positive” with no distinction in terms of the type of violence and sentences with no violence are labeled as “negative” (including related and unrelated negatives).

We also consider all positive and negative testing examples in testing, with no distinction with respect to type. The results of this analysis are shown in Figure 2.

**Figure 2:**
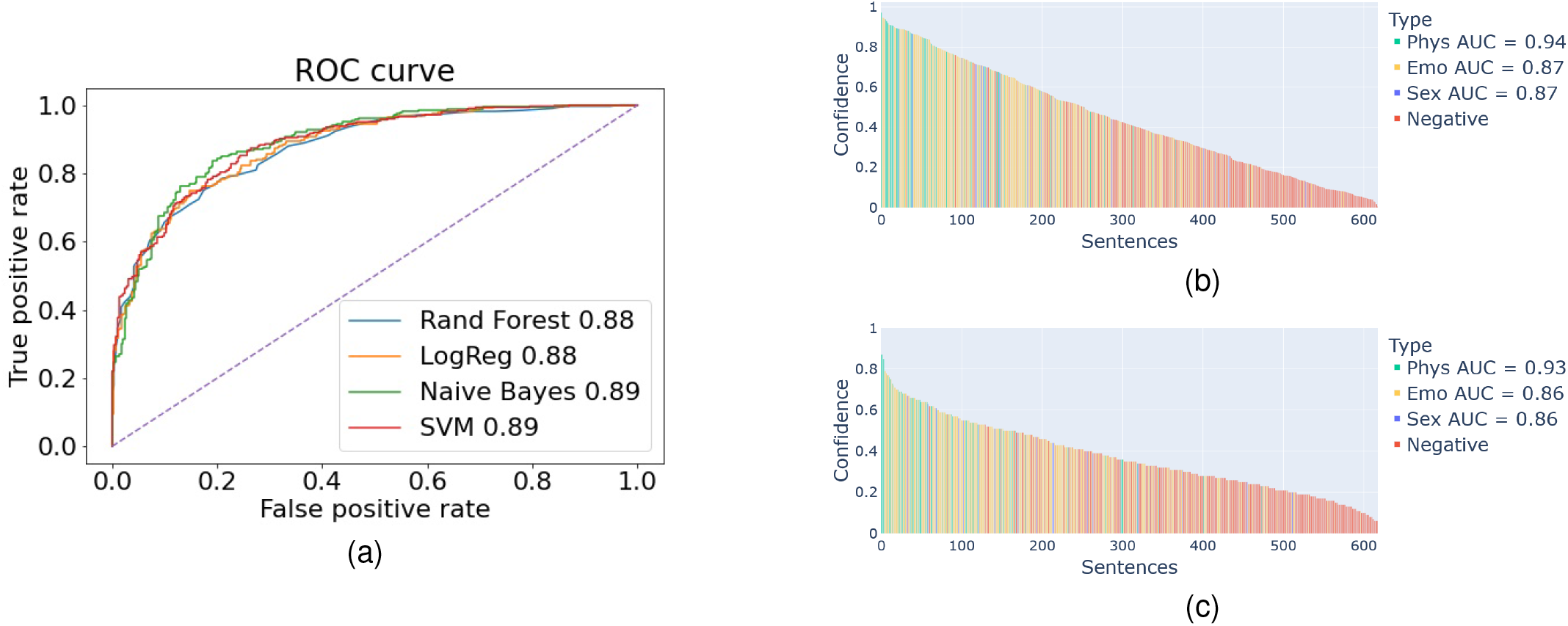
Predictive accuracy of the “General Violence Model” using different machine learning algorithms. (a) The ROC curve for the predictions provided by four different machine learning algorithms on the testing (social media) data, where sentences are labeled “positive” or “negative” depending on the presence of violence in the sentence. (b) The predictions of the Logistic Regression model on the testing (social media) data, where sentences are ordered according to confidence of predictions and colors show sentence label such that positive samples are annotated with type of violence. (c) The predictions of the Random Forest model on the testing (social media) data. Phys: Physical Abuse, Emo: Emotional Abuse, Sex: Sexual Abuse, Negative: No Violence. Each violence type is indicated with Area Under ROC, or Area Under Curve (AUC)

Overall, we observe that the models built using four different classification algorithms deliver similar predictive accuracy, where the area under ROC curve (AUROC) is slightly less than 90% for all algorithms. The waterfall plots provide further insights into the predictions of Logistic Regression and Random Forest Models, in which the positive testing examples are colored according to violence type. In these waterfall plots, the test sentences are ordered on the x-axis according to the confidence assigned by the classifier in labeling the sentence as “violence” and this confidence value is shown on the y-axis. As seen in these plots, both classifiers are able to assign high confidence scores to sentences with Physical Abuse, while confusion mostly occurs between sentences that contain Emotional Abuse and negatives. We also observe that the Logistic Regression Model is slightly more successful in distinguishing Sexual Abuse from negatives.

### 3.2 Predictive Performance of Type-Specific Violence Models

As the results shown in the previous section demonstrate, the General Violence model provides nearly 90% AUROC in detecting IPV in a sentence. When we consider the accuracy of this model in detecting specific types of violence, we observe that it is most successful in bringing forward Physical Abuse, while its performance is variable in detecting Emotional or Sexual Abuse. This observation leads to the question of whether a type-specific model that is trained using only the specific violence type for positive examples can provide better performance in detecting the target violence type. To answer this question, we compare the performance of the Type-Specific Violence Model in detecting their target violence type against that of the General Violence Model. The results of this analysis using Logistic Regression are shown in Figure 3.

**Figure 3:**
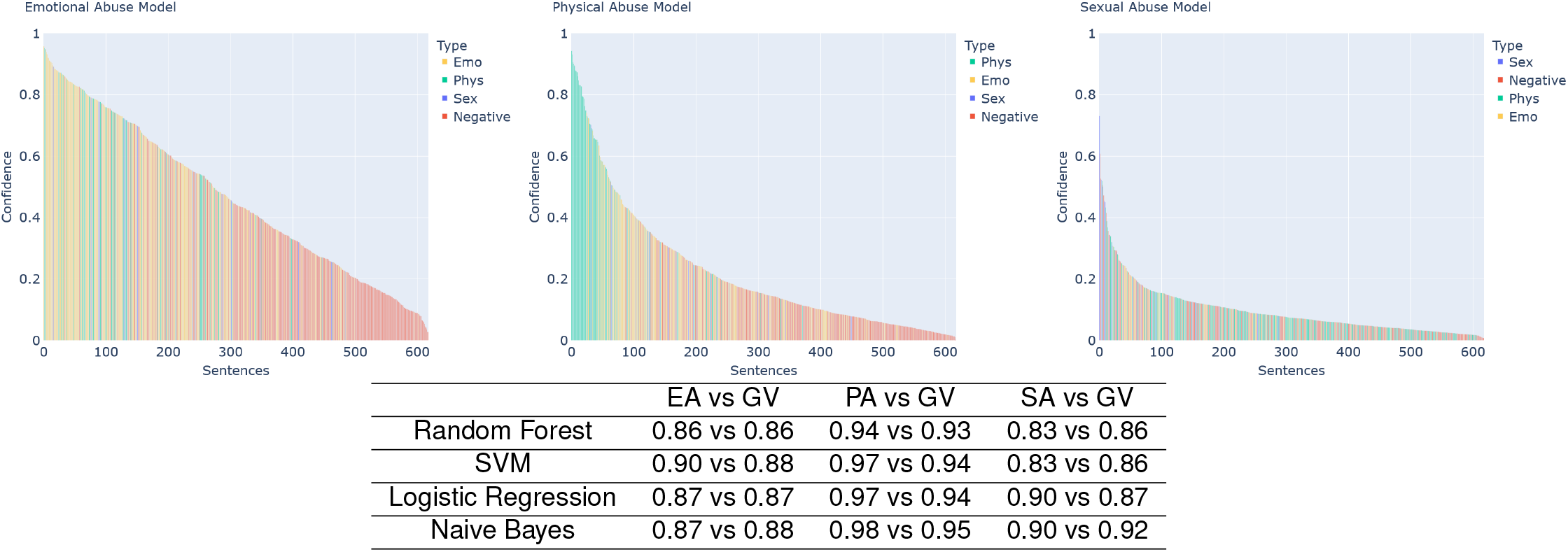
Predictive performance of type-specific violence models. The type-specific models are trained by considering the specified IPV types as positive samples and sentences with no violence as negative samples. For a type-specific model, positive samples representing the other two types are not included in the training data. Upper panel: Waterfall plot of the confidence scores assigned by the type-specific model to all sentences in the test (social media) data for Emotional, Physical, and Sexual Abuse models from left to right. The bars are colored according to violence type (or no violence/negative). Lower panel: The comparison of the performance of the type-specific violence model in predicting violence against the general violence model based on all type-specific sentences and negatives in the test (social media) data (type-specific (left) vs general (right)). Positive test samples representing the other two types are not consider in computing the ROC.

As seen in Figure 3, the type-specific Physical Abuse model performs even better than the General Violence Model in detecting Physical Abuse. This is also true for Sexual Abuse, where the type-specific model can detect Sexual Abuse with 90% AUROC. In contrast, the type-specific model for Emotional Abuse does not improve the detection of Emotional Abuse model as compared to the general model that also makes use of examples of Physical and Sexual Abuse. Of particular note is the shape of the waterfall plots for each of the type-specific models. While the EA-specific model distributes confidence scores uniformly across all test sentences, the PA-specific scores sentences with physical abuse with very high confidence, while assigning considerably low confidence scores to a majority of the test sentences (most of which are true negatives). The SA-specific model, on the other hand, tends to assign lower confidence scores to all test sentences -this is likely due to the relative scarcity of sentences with sexual abuse in our training and test data.

### 3.3 Visualization of Sentences in the Space of Violence Types

To gain further insights into what the algorithms learn to detect IPV in sentences, we visualize the predictions of the three type-specific violence models in the 3-dimensional space of violence types. For this purpose, we use the radial visualization technique developed by Hacialiefendioğlu et al. ^11^. The results of this analysis for Random Forest and Logistic Regression models are shown in Figure 4. In the figure, the predictions of the models for training sentences are visualized on the left. For these sentences, the relative location of each point (representing a sentence) shows how much the model is able to fit the sentence to its true label (since these sentences are used in training). The web service implementation of DetectIPV visualizes the query sentences in this radial space, providing users with the ability to visually explore the confidence of the models for each sentence.

**Figure 4:**
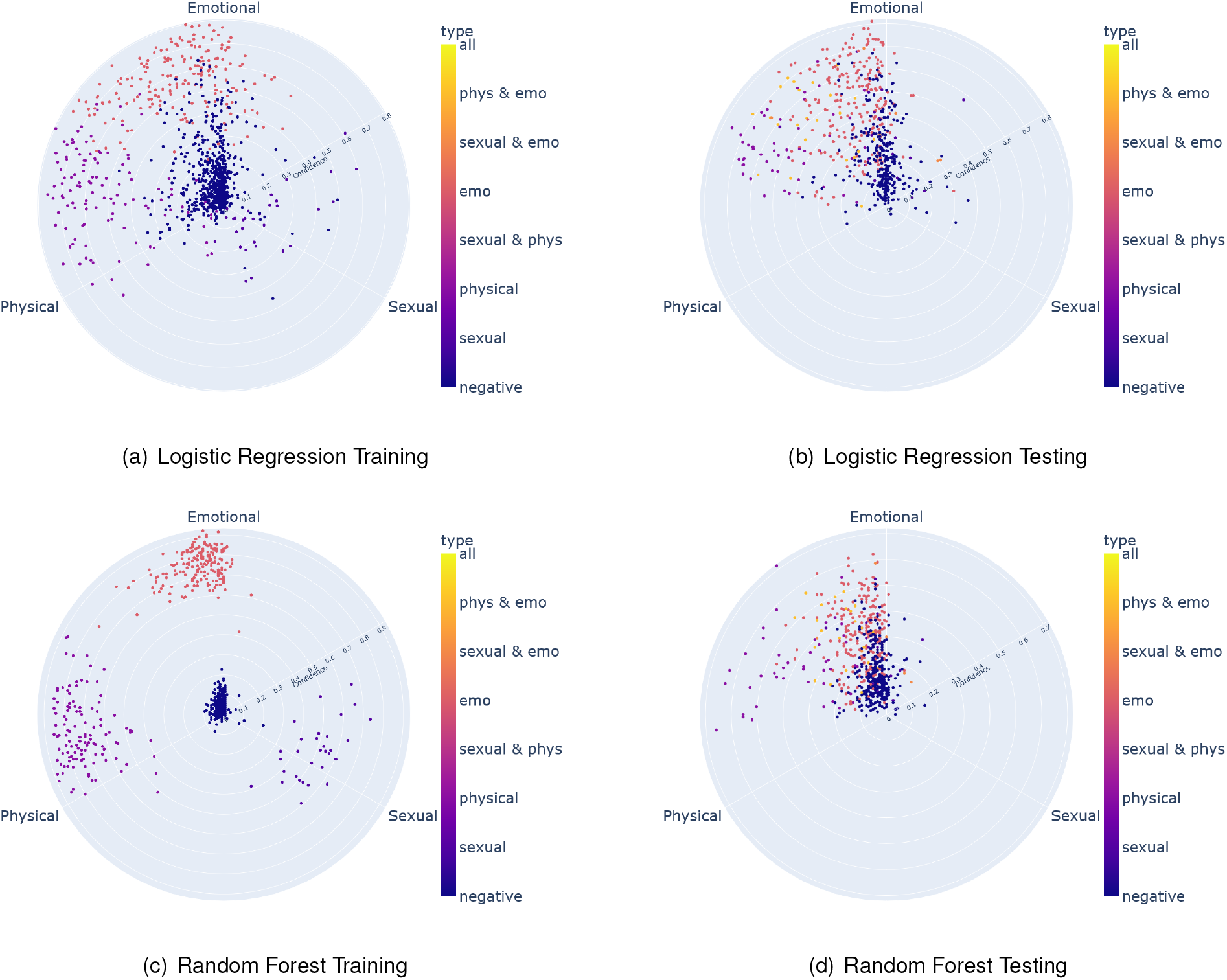
Radial visualization of the predictions of type-specific models for training and testing sentences. Upper panel: Random Forest model, Lower panel: Logistic Regression model; Left: Training (Questionnaire) Sentences, Right: Testing (Social Media) Sentences. Each axis represents an IPV type (0°: Emotional Abuse, 120°: Sexual Abuse, 240°: Physical Abuse). The distance of a point from the center represents the magnitude of the degree of confidence assigned by the three type-specific models to the corresponding sentence, while its angle/direction represents the dominant violence type in terms of the confidence assigned by the models. The color of the point represents the true label assigned to each sentence by expert reviewers, which can include of multiple types of IPV, as shown by the color bar.

As seen in Figure 4, the Random Forest model is able to fit the data in a way that clearly separates different types of samples in the space of violence types. Logistic Regression, on the other hand, incorporates the overlap between Physical and Emotional Abuse, as well as the confusion between negative sentences and Emotional Abuse into the model. Interestingly, however, as shown on the right panel of the figures, the two models behave similarly on test sentences. Namely, the models can clearly separate Physical Abuse from other types of violence (or negatives), while there is a lot of overlap between Physical and Emotional Abuse predictions, negative sentences are mostly confused with Emotional Abuse, and the models assign low confidence to Sexual Abuse in general.

### 3.4 Comparison Against Alternate Approaches and Effect of Number of Dimensions

To understand the benefits of using sentence embedding, we compare the predictive accuracy of DetectIPV against a term-frequency based approach, in this case, TF-IDF with single words and bi-grams. While doing so, we also investigate the effect of the number of dimensions/features used for classification. For Universal Sentence Encoder, the features are the dimensions in the embedding space. To prioritize and select these features, we use a filtering approach using the separation of labeled training (questionnaire) sentences, as described in section 2. For the term-frequency based approach, the features are words and bigrams. To prioritize and select the term-frequency based features, we compute the mutual information between the presence of each term with sentence labels in the training (questionnaire) data and select features in decreasing order of mutual information. The results of this analysis are shown in Figure 5.

**Figure 5:**
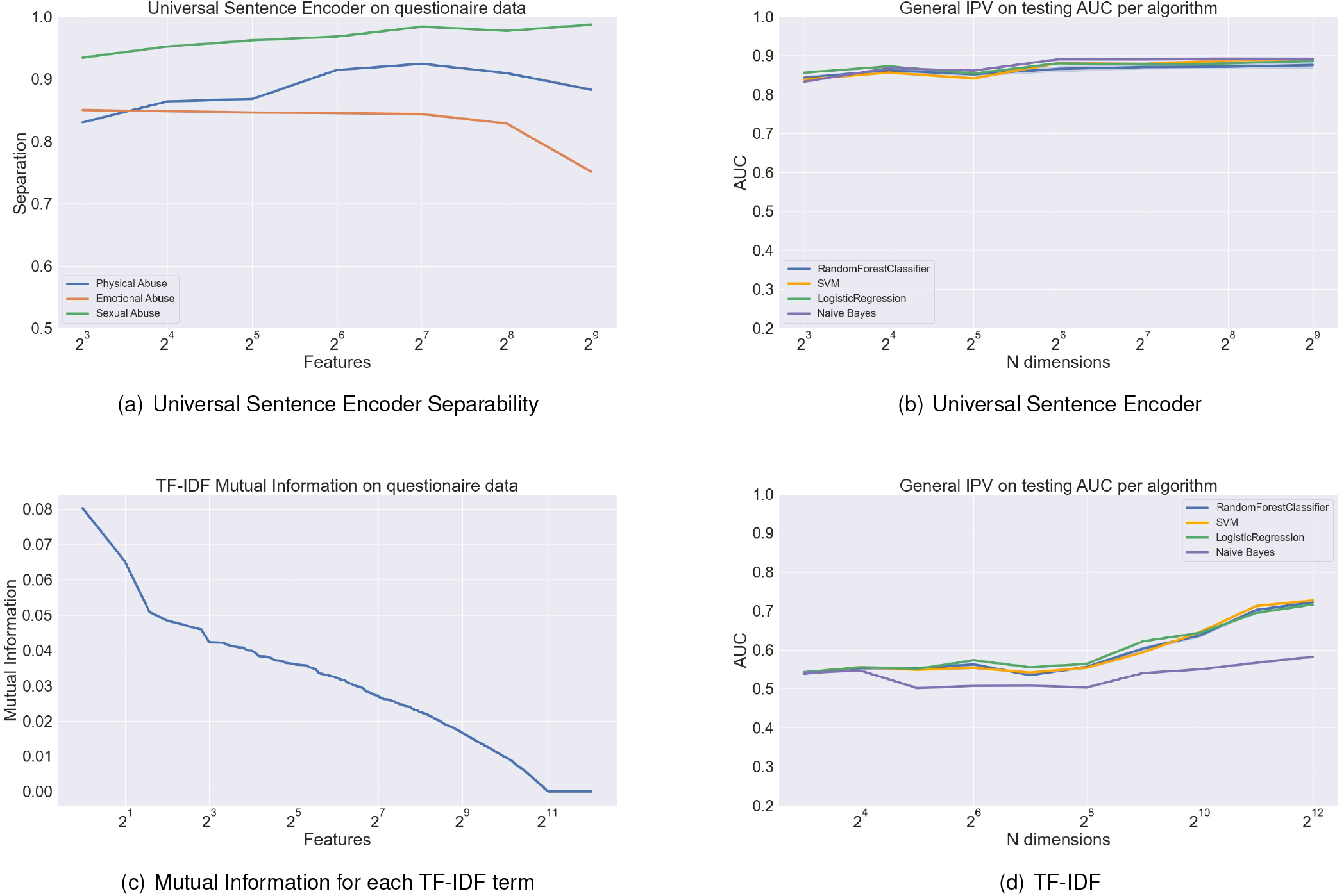
Comparison between Universal Sentence Encoder (USE) vs. term-frequency (TF-IDF) based prediction of IPV. Left: Feature prioritization and selection. For USE, dimensions are ordered according to separability of general violence, cumulative separability of each violence type is shown in the plots (a). For term-frequency, the terms are ordered according to mutual information with sentence labels and rank vs. mutual information is plotted in (c). Right: The predictive performance (AUROC) of the general violence models on the test (social media) sentences as a function of number of features using USE (b) and TF-IDF (d). The features are selected based on the ordering on the left (computed on training sentences) and four different models are trained for each number of features using four different classification algorithms.

As seen in Figure 5, the predictive performance of DetectIPV goes up with increasing number of USE dimensions, and stabilizes at around 128 dimensions. This is also captured by our measure of separability on the training data, the separability of physical and emotional violence peaks at around 128 dimensions as well. Importantly, all four classification algorithms perform fairly similarly in detecting general violence when presented with the same set of features. It is interesting to note that these models perform fairly well (above 80% AUROC) even with the first 8 dimensions dimension, suggesting that the first 8 dimensions selected from USE provides a representative latent feature for the presence of violence in a sentence.

In contrast to DetectIPV, the TF-IDF models perform sligthly better than random (around 50% AUROC) when they utilize a low number of dimensions (terms), although the first few terms are most informative on violence as quantified by mutual information. As the number of terms go up, the predictive accuracy of TF-IDF models improves, and nears 75% AUROC when all words and bigrams are used, except for the Naïve Bayes model. Overall, these results show that the use of sentence embeddings in clearly outperforms TF-IDF based models in detecting violence in sentences.

### 3.5 The Effect of Negative Examples Used in Training

As discussed in Section 2, we use negative examples that are unrelated to relationships, as well as those that are on relationships, but do not include violence or abuse. Since DetectIPV potentially has a broad range of application domains, one can be interested in using it to identify violence in a corpus on relationship problems or identify IPV-related sentences in a larger, broader corpus. Motivated by this consideration, we comparehensively characterize the predictive accuracy of DetectIPV in distinguishing IPV-related sentences from sentences that are related or unrelated to relationships. The results of this analysis are shown in Table 3.

**Table 3:** The effect of using relationship-related or unrelated negative sentences in training and testing on predictive performance. Each entry shows the area under ROC curve for four machine learning algorithms on a task obtained by using relationship-related negatives, unrelated negatives, and both types of negatives in training vs. testing. The numbers for each machine learning algorithm is shown in a different color: Random Forest (cyan), SVM (orange), Naive Bayes (purple), Logistic Regression (olive)

| Negative Type | Testing Unrelated |  | Testing Related |  | Testing Both |  |
| --- | --- | --- | --- | --- | --- | --- |
| Training Unrelated | 0.91 | 0.93 | 0.77 | 0.81 | 0.86 | 0.89 |
|  | 0.93 | 0.92 | 0.78 | 0.81 | 0.88 | 0.88 |
| Training Related | 0.83 | 0.89 | 0.78 | 0.82 | 0.82 | 0.86 |
|  | 0.90 | 0.87 | 0.79 | 0.80 | 0.86 | 0.85 |
| Training Both | 0.91 | 0.91 | 0.83 | 0.84 | 0.88 | 0.89 |
|  | 0.94 | 0.91 | 0.80 | 0.82 | 0.89 | 0.88 |

As would be expected, DetectIPV delivers best predictive performance (above 90% AUROC for all models) in distinguishing IPV-related sentences from unrelated sentences when trained using unrelated negative examples. These models’ AUROC goes down to nearly 80% when they are tested on a set that includes only relationship-related sentences as negatives. This observation suggests that models trained using unrelated negatives learn to detect presence of violence, but they also learn to detect relationship issues to a certain extent. However, training on relationship-related sentences does not significantly improve the performance of this algorithms in distingushing IPV-related sentences from relationship-related sentences (AUROC around 80% for all models). Importantly, in distinguishing IPV-related sentences from unrelated sentences, the four models that are trained using both related and unrelated negatives perform nearly as good as models that are trained only using unrelated negatives. These results show that DetectIPV is quite robust to the types of sentences that are used in training and testing, and almost always delivers nearly 80% accuracy. These results also provide insights into selecting negative examples to train DetectIPV depending on the application.

## 4 Discussions

IPV has many devastating consequences on the victim’s emotional and physical well being with substantial morbidity and mortality. IPV does not only involve tangible violence but also involve abusive situations that are harmful to emotional well being. These abusive situations can often be unrecognized and under-reported ^79 80^ Our results show that the general model that is trained using examples of all violence types can identify IPV from free text with high (*>* 90%) confidence. We also train type-specific models and observe that Physical Abuse can be identified with high precision, while Sexual Abuse can be identified with high precision but relatively low recall. Our results also indicate that the prediction of Emotional Abuse is the most challenging.

Our model can distinguish Physical Abuse more distinctly as compared to other violence types. It is possible that signs of Physical Abuse are more noticeable. It is usually more clearly indicated as the language used to describe Physical Abuse is usually quite specific, such as hitting, scratching, pushing, shoving, throwing, grabbing, choking, biting, hair-pulling, slapping, hitting, punching, burning or use of restraints/body size and/or strength against another person. Furthermore, Physical Abuse is usually a patterned behavior in that it is usually not an isolated incident, but rather becomes gradually more frequent and also co-occurs with Emotional Abuse.

Our results indicate that Emotional Abuse is the hardest type of violence to distinguish among the free text. Emotional Abuse, the continual emotional mistreatment of a person, diminishes a person’s self-worth or emotional well-being and is associated with the development of depression, anxiety and post-traumatic stress disorder (PTSD) ^81^. Emotional Abuse is also considered a precursor to Physical and Sexual Abuse. Identifying and interpreting Emotional Abuse presents a challenge due to the broad range of behaviors that may fall into this category and the need for contextualization.

Emotional Abuse is often not visible, it can be passed and unnoticed in conversation or posts. It is often harder to identify indicators of Emotional Abuse online as the tone of the post is not as easily understood. Those who are not aware of their situation will change the language they use when describing their experience. This may be confusing to our model and can trigger false positives or false negatives. However, certain words can imply a level of Emotional Abuse including but not limited to threat/threatened, control, fear/afraid, manipulate, neglect, demean, gaslit/gaslighting. In addition, the use of semantic embedding (Universal Sentence Encoder) can help capture context to a certain extent. Thus the sentence-focused approach of DetectIPV is able to detect Emotional Abuse with reasonable and potentially useful accuracy. However, approaches that use broader context (e.g., multiple sentences) and more sophisticated nature-language processing (NLP) algorithms can improve the accuracy of detecting Emotional Abuse in free text.

Our results indicate that when distinguishing Sexual Abuse, results indicate the importance of looking at a wider range of key terms, which may not be individually associated with violence. In general, sentences involving Sexual Abuse indicate both the physical and emotional nature of Sexual Abuse in intimate relationships.

The threat to violence and verbalizing these threats could be warning signs for severe and dangerous violence. Campbell’s ^20^ danger assessment survey indicates that such threats often increase the risk of danger and homicide. Threats may also happen simultaneously with Emotional, Sexual, and Physical Abuse. It is possible that our model might incorrectly label threats as one or another form of violence. Similarly, although considered as Emotional Abuse, property damage can also involve physical violence as it can be viewed as symbolic violence and comfort with destructive power. ^82^ Relationships involving threats of violence and property damage can be labeled more accurately utilizing context-dependent information.

### Limitations

An important limitation of this study is the scale of data that is available for training and testing. Specifically, for violence types, training data was more scarse for Sexual Abuse, due to the availability of a limited number of measures with good psychometric qualities. An important factor that contributes to the accuracy of machine learning models is the size of training data. Here, by (i) using training and testing examples that come from different sources and (ii) utilizing pretrained semantic embeddings to represent sentences, we alleviated the effect of relatively small sample size to a certain extent. However, availability of abundant data will enable training of more sophisticated machine learning models (e.g., deep learning) and likely enhance accuracy of predictions. Nevertheless, this study represents a significant first step towards development of such large-scale machine learning models, by providing insights into the effect of many factors, including violence types, negative examples used in training, and features that are used. Importantly, DetectIPV can also be used to generate a large corpus of text describing IPV, by iteratively applying DetectIPV to prioritize sentences and reviewing texts that are enriched in sentences scored highly by DetectIPV.

The test data that is collected from the social media may also be biased in various ways, which may influence the conclusions of this study. It is possible that people who are in abusive relationships, particularly for specific types or severity of abuse, may not post for advice publicly due to stigma and more serious red flags. This may bias the test data toward specific types of violence and/or levels of severity. In addition, red flags can be less noticeable to the victim, as Physical Abuse tends to be progressive over time than a sudden outburst. Thus it can be difficult to find Emotional Abuse in a post possibly commented in a nuanced way in the main text. IPV also has many unexplored comorbidities. In future research, our investigation of relationship dynamics that are significantly different from violent and not violent relationships can investigate the situational and context-dependent nature of IPV.

## 5 Conclusion

IPV can adversely effect mental, sexual and physical health. Not being able to detect signs of red flags can danger the safety of the individuals. Early and accurate characterization of IPV can help the victims navigate their situation to relieve certain emotional, sexual and physical consequences and guide treatment for the victims. Providing tools for exploring potential issues in their relationships is critical for improving their health and safety.

The primary outcome of this study is DetectIPV, a tool for detecting IPV from free text. DetectIPV is carefully developed and validated by taking into account multiple factors. It is available as a web service that can be used to detect IPV in query texts using pre-trained models, as well as open-source code that can be trained using other sets of training data and used as a stand-alone tool. DetectIPV can be utilized in a broad range of applications, including flagging IPV in social media, detecting IPV in electronic health records, detecting IPV in court records, detecting IPV in transcripts of interviews, and assessing IPV in large corpuses of text for research purposes.

## Data Availability

All data produced in the present study are available upon reasonable request to the authors

## 6 Acknowledgement

This publication was made possible by US National Health Institutes (NIH) grant R01-LM012518 from the National Library of Medicine. Its contents are solely the responsibility of the authors and do not necessarily represent the official views of the NIH.

## Confilct of Interests Statement

On behalf of all authors, the corresponding author states that there is no conflict of interest.

## Notes

### Competing Interest Statement

The authors have declared no competing interest.

### Funding Statement

This study was funded by US National Health Institutes (NIH) grant R01-LM012518 from the National Library of Medicine

